# Predicting SARS-CoV-2 infection trend using technical analysis indicators

**DOI:** 10.1101/2020.05.13.20100784

**Authors:** Marino Paroli, Maria Isabella Sirinian

## Abstract

COVID-19 pandemic is a global emergency caused by SARS-CoV-2 infection. Without efficacious antiviral drugs or vaccines available, mass quarantine has been the main strategy adopted by governments to contain the virus spread. This has led to a significant reduction in the number of infected people and deaths and to a diminished pressure over the health care system. However, an economic depression is following due to the forced absence of worker from their job and to the closure of many productive activities. For these reasons, governments are lessening progressively the mass quarantine measures to avoid an economic catastrophe. However, most people has not been yet exposed to the virus, so are susceptible to Covid-19. Therefore, the reopening of firms and commercial activities might lead to a resurgence of infection, as already reported in South Korea and in China were epidemic was considered almost gone although in a few of cases. In the worst-case scenario, this might impose the return to strict lockdown measures. Epidemiological models are therefore necessary to forecast possible new infection outbreaks and to inform government to promptly adopt new containment measures. In this context, we tested here if technical analysis methods commonly used in the financial market might provide early signal of change in the direction of SARS-Cov-2 infection trend in Italy, Iran and Brazil, three countries which have been strongly hit by the pandemic but whose present infection trends are substantially different from each other being therefore particularly useful to understand the potential of TA models. We conclude that technical analysis indicators can be usefully adopted to this aim.

## INTRODUCTION

Severe Acute Respiratory Syndrome Coronavirus 2 (SARS-CoV-2) is the etiological agent of Coronavirus Disease 2019 (COVID-19) (1). At the time of writing, COVID-19 pandemic is a global emergency with more than 280,000 deaths worldwide (https://data.europa.eu/euodp/en/home). With no effective antiviral drugs and no vaccines available, prevention of COVID-19 relies upon detection and isolation of symptomatic cases and restrictive mass quarantines (2). Mass quarantine, however, poses a serious risk of a second crisis in the form of an economic recession (3). Therefore, governments are cautiously allowing the progressive come back to work without certainty that further infection waves will not occur. Different epidemiological models are used to forecast real-time the number of new cases and identify possible pandemic outbreaks. These include compartmental models, agent-based models and metapopulation models (4-5). However, any epidemiological model has its own limitation, and the addition of new predictive tools is desired. Technical analysis (TA) is a methodology used in the financial market aimed to forecast the direction of security prices through the study of statistical trends of post-market data (6). TA is based on the use of mathematical tools called indicators developed to generate signals for traders to buy or to sell. These tools include indicators just plotted over the top of the prices on a stock chart like Simple Moving Averages (SMEs) or which oscillate between minimum and maximum value like Moving Average Convergence/Divergence (MACD) and the Relative Strength Index (RSI) (7-8). We examined here the capacity of different TA indicators to predict the SARS-CoV-2 spreading using daily reported new cases of infection for our data analysis.

## METHODS

Open data of new daily infection cases in Italy, Iran and Brazil were obtained from European Union official website (9). TA indicators used in this study included a) the combined use of fast (3-day) and slow (20-day) Simple Mean Averages (SMAs). In particular, the crossing between the fast and the slow SMAs either to the upside or the downside are considered as signs of trend reversals; b) Moving Average Convergence Divergence (MACD). MACD is a commonly used indicator which consists of the so-called MACD line calculated by subtracting the 26-day Exponential Moving Average (EMA) from the 12-day EMA of a series of consecutive data. Nine-day EMA of the MACD line (signal line) is then calculated and plotted on top of the MACD line. Finally, MACD histogram is calculated by subtracting the signal line from the MACD line. A trend reversal to the upside is signaled when the MACD line crosses above its signal line, while a reversal trend to the downside is signaled when the MACD line crosses below the signal line. Additionally, signals of a trend reversals are revealed when the MACD line cross the zero axis; c) the 14-day Relative Strength Index (RSI) was calculated according to the standard formula: RSI = 100 − (100/1+ Relative Strength (RS)) where RS is the ratio between the average of the absolute increase of values upper and down the previous day in a 14-day timeframe. Plotting RSI values on a chart results in a line that fluctuates between 0 and 100 values. The crossing of the 50 value by the RSI line either to the upside or to the downside are considered reversal signals of the current trend; f) divergence between MACD histogram and daily new infections trends were also analyzed. All TA indicators were calculated according to the appropriate formula using the Microsoft Excel spreadsheet program.

## RESULTS

Trend analysis of daily new infections in Italy (fig. 1A) showed that the fast (3-days) SMA crossed the slow (20-days) line from above on April 1, 2020 indicating that the upward trend was likely to change in a downward direction. This signal occurred about 10 days after that a peak in the number of new infected cases was recorded on 22 March, 2020. MACD analysis (fig. 2A) showed to subsequent signals or trend reversal to the downside. The first signal consisting in the crossing of MACD line by the signal line from above appeared on 30 March and the second one on 20 April. Finally, RSI (Fig 3A) showed the crossing of 50-axos by the RSI line. In Iran (fig. 1B) a signal of reversal to the uptrend appeared on April 8, about 8 days after the highest number of newly reported cases on March 31 (solid arrow). Thereafter, and uptrend signal followed on May 6 represented by the crossing of slow SMA by the fast SMA from above (dotted arrow). Similarly, MACD analysis signaled two opposite trend reversal signals. The first was a downtrend signal and appeared on March 24 followed by an uptrend signal on April, 5. This trend was confirmed by the zero-axis crossing by MACD line on April 20. A new trend reversal appeared on May 5 concomitant with the change of MACD histogram from negative to positive. After this signal, the trend reversal was confirmed by an increase of the daily reported infections. In fig. 3B the RSI line signaled a trend inversion to the downside on March 28 and to the upside on April 24 by crossing the 50-axis. As regards Brazil (fig. 1C, 2C and 3C) the trend of new infections was constantly toward the upside and no signal of reversal was signaled during all the timeframe observed. Finally, divergences between trends (dotted lines) of the number of new daily cases (solid line) and MACD (histograms) were alalyzed. In Italy (fig. 4A) a divergence appeared during the downtrend phase of the epidemic, as indicated by the dotted lines. However, no trend reversal occurred so this should be considered as a false signal. In Iran (fig 4B) the chart displayed two divergences. The first one was noticeable in the early phase of the time period examined. This also was a false signal. After the second divergence, however, a trend reversal actually occurred (dotted arrow). In Brazil (fig. 4C) no trend divergence between case of infections and MACD histogram was found.

**Figure 1.**
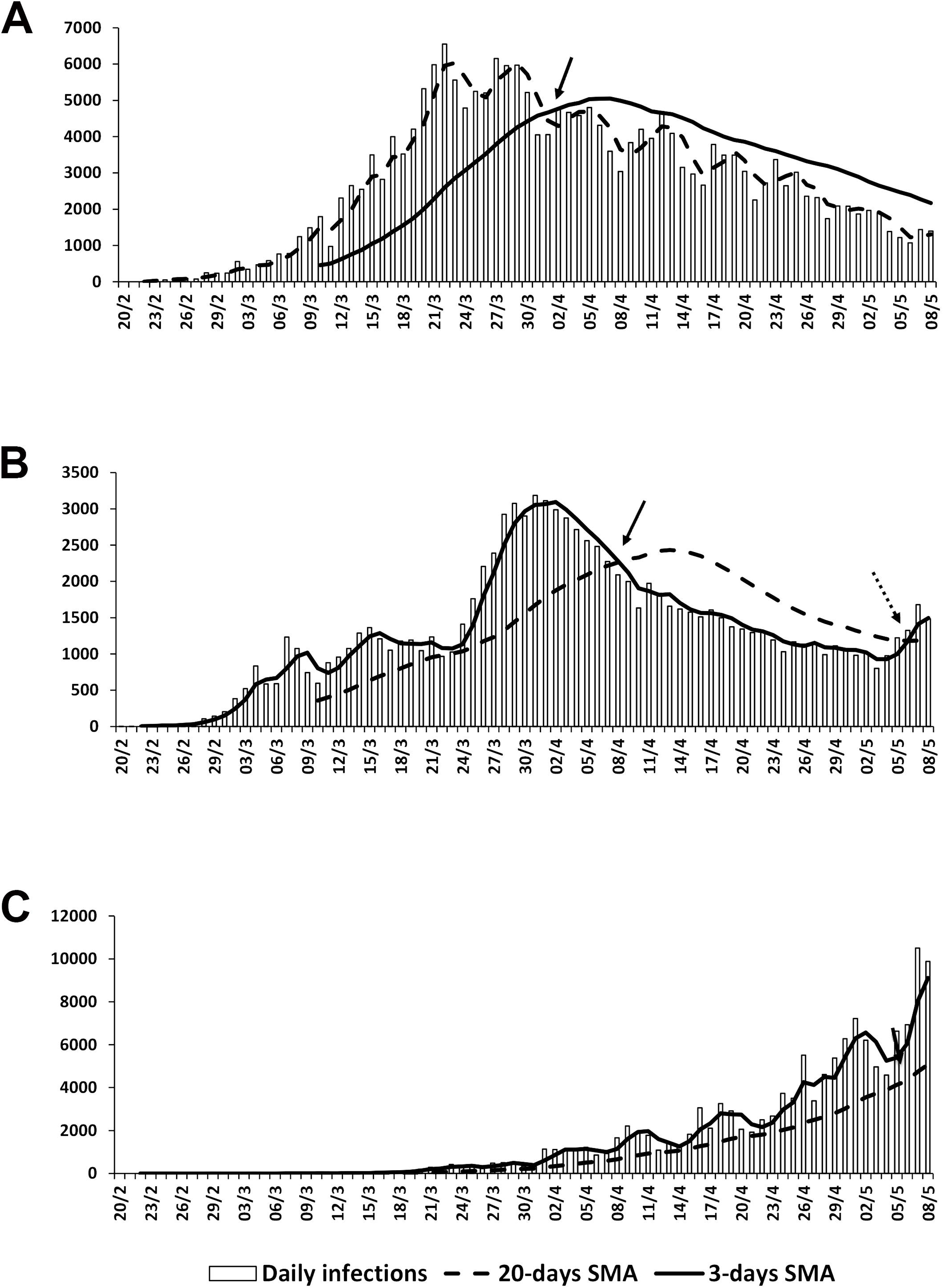
In Italy (A) the fast SMA crosses the slow SMA from above (red arrow) indicating that the trend is changing in a downward direction. Histograms represent the reported new daily infections. In Iran (B) a first signal of trend reversal to the downside (red arrow) is followed by a trend reversal to the upside (black arrow). In Brazil (C) the upward trend is constant without any signals of reversal.

**Figure 2.**
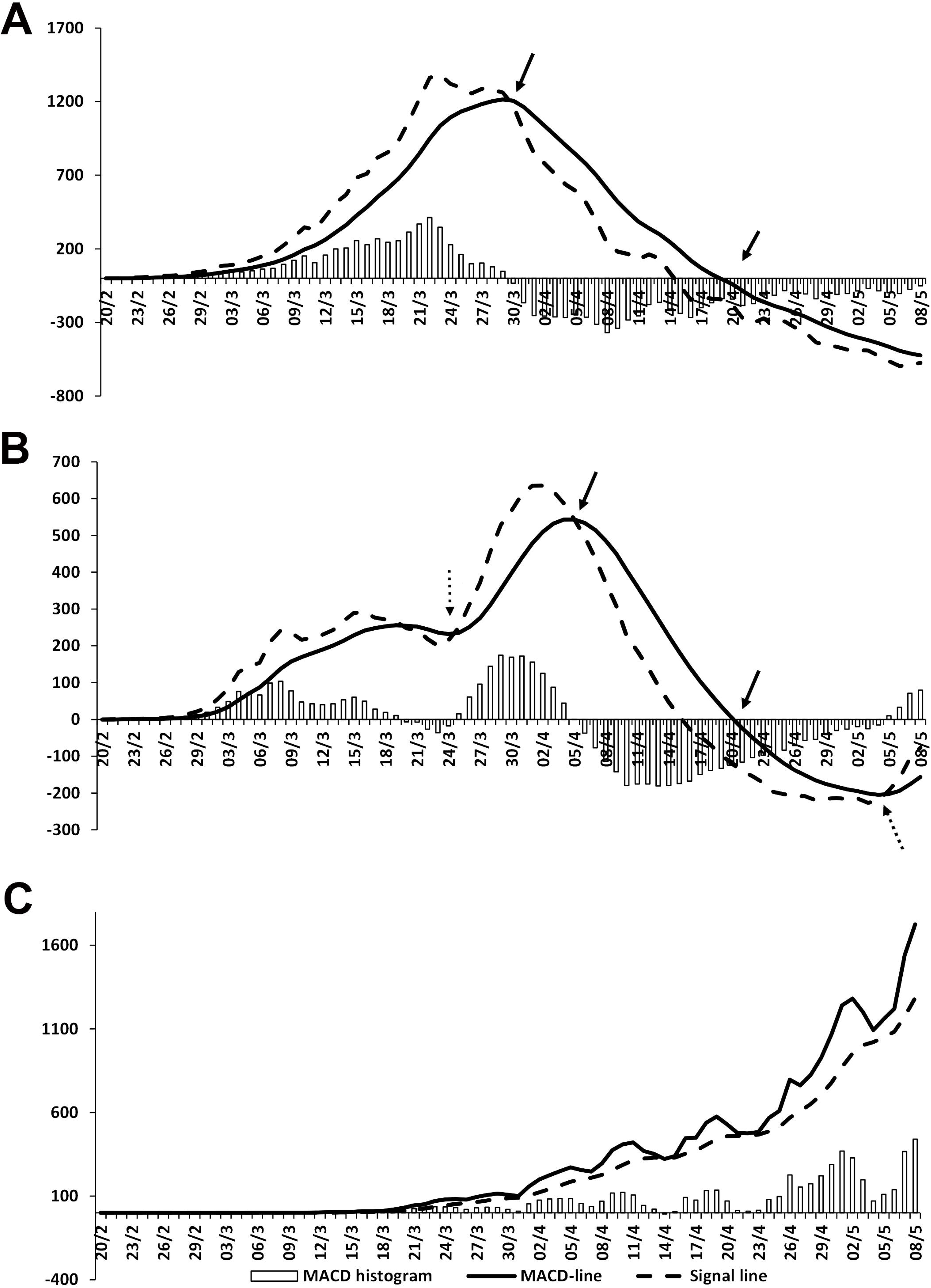
A trend reversal to the downside in Italy (A) is first signaled by the crossing of the faster signal line to the slower MACD line from the above and confirmed later by the crossing of the zero-axis by the MACD line from the above (red arrows). In Iran (B) the red arrow signals an initial downtrend followed by a return to the upside as signaled by the black arrow. In Brazil (C) no signal of trend reversal is spotted.

**Figure 3.**
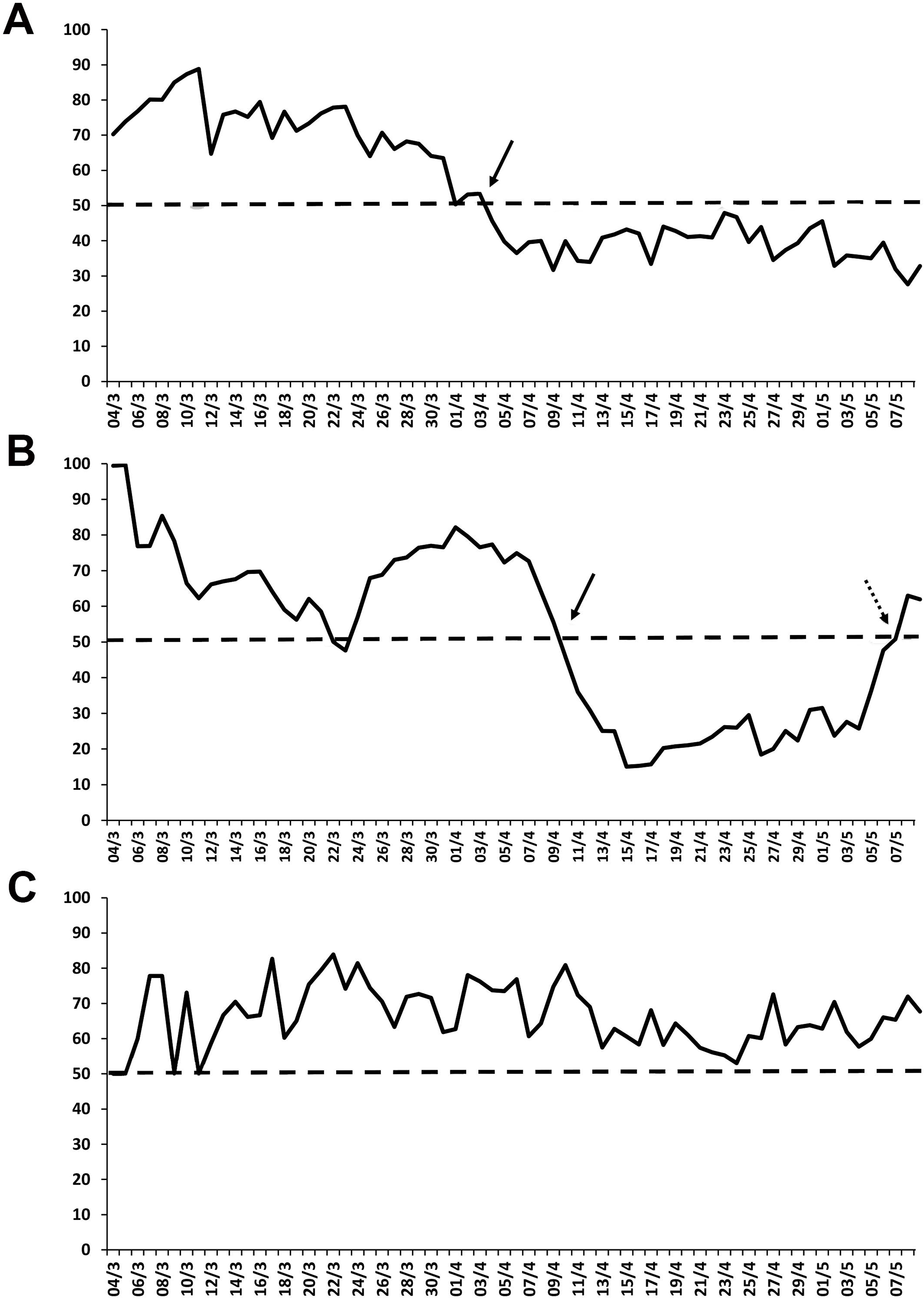
Relative strength indicator (RSI) is shown. In Italy (A) the crossing of the 50-line by the RSI line confirms a trend reversal to the downside (red arrow). In Iran (B) a signal of reversal to the uptrend follows a first signal to the downtrend (red and black arrows, respectively). In Brazil (C) no signal of trend reversal is detected.

**Figure 4.**
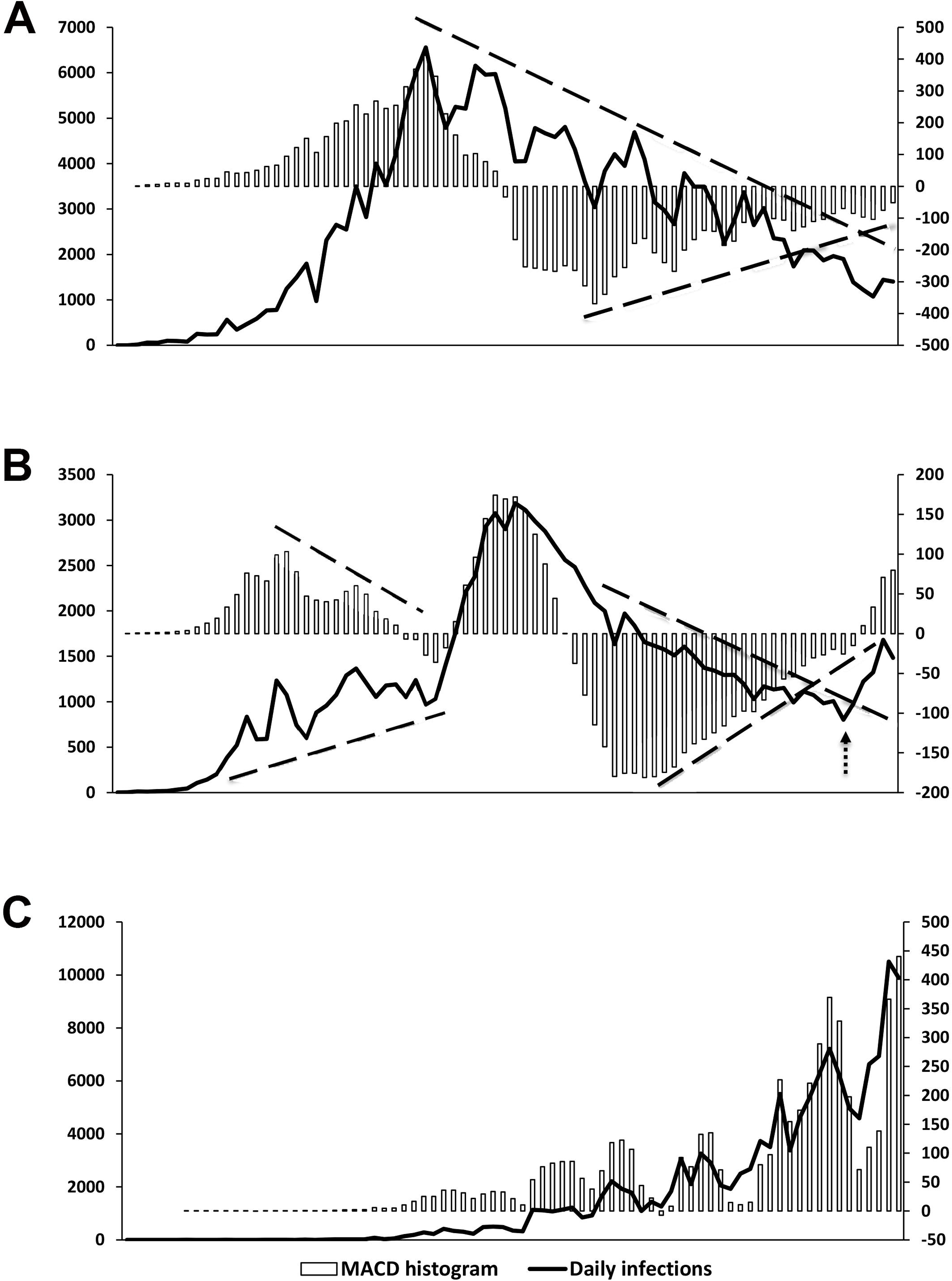
Divergences between the number of new daily cases (solid line) and MACD (histogram) are shown. In Italy (A) a divergence appears during the downtrend phase of the epidemic, as indicated by the dotted lines. However, no trend reversal occurs (false signal). In Iran (B) two divergences occur at the first and second phases of the epidemic. After the second divergence, a trend reversal actually occurs (black arrow). In Brazil (C) no divergence is found.

## DISCUSSION

Technical analysis is a means to examine and forecast how financial market will trend. TA is based on the idea that if previous market patterns can be identified a fairly accurate prediction of future price trajectories can be made, and this is obtained by the use of mathematical tools called market indicators. This approach is opposed to fundamental analysis which is focuses on the true values of assets (6). A series of TA indicators were tested here for their ability to predict SARS-Cov-2 infection spread using as data the number of daily reported new infections in analogy to stock quote prices. Open data on infections obtained by the European official site concerned Italy, Iran and Brazil, all countries greatly struck by the pandemic. We found that all indicators considered here provided trend reversal signals which were easily spotted on charts. This led us to the conclusion that TA indicators can be useful to obtain real-time information on how SARS-CoV-2 infection is spreading. It is worth to note that prompt signals of a trend inversion can provide essential information to governments to adopt prompt measures to contain the pandemic and to identify new outbreaks. Although TA is not intended to be a substitute of classical epidemiological models, we believe that any new method able to predict the infection trend deserve consideration. This is true especially during such an emergency situation where at present neither the biology of the virus nor the host immunological response is fully understood. A major limitation of this study is the intrinsic nature of the data. The reported number of daily new cases is at least in part inaccurate since a consistent number of new infections can be missed due to the presence of asymptomatic subjects (10). Underreporting of cases due to several factors is a major limitation of any epidemiological model. Moreover, the number of diagnostic tests taken daily may fluctuate significantly. It is worth to note that the number of tests does not refer to the same in each country. One main difference is that some countries report the number of people tested, while others report the number of tests which can be higher if the same person is tested more than once. Finally, the incidence of infection can vary among areas of the same country not necessarily reflecting the situation of the whole country (11). This has been particularly evident in Italy where in Lombardy region cases account for about half of total cases of Italy. Nevertheless, we suggest that TA indicators might provide reliable real-time information of how SARS-Cov-2 infection is spreading to rush the set-up of new containment measures by governments when needed.

## Data Availability

Data have been obtained from official EU website

https://data.europa.eu/euodp/en/home

